# The Canadian ALS Neuroimaging Consortium (CALSNIC) - a multicentre platform for standardized imaging and clinical studies in ALS

**DOI:** 10.1101/2020.07.10.20142679

**Authors:** Sanjay Kalra, Muhammad Khan, Laura Barlow, Christian Beaulieu, Michael Benatar, Hannah Briemberg, Sneha Chenji, Miriam Garrido Clua, Sumit Das, Annie Dionne, Nicolas Dupré, Derek Emery, Dean Eurich, Richard Frayne, Angela Genge, Summer Gibson, Simon Graham, Christopher Hanstock, Abdullah Ishaque, Jeffrey T. Joseph, Julia Keith, Lawrence Korngut, Dennell Krebs, Cheryl R. McCreary, Pradip Pattany, Peter Seres, Christen Shoesmith, Trevor Szekeres, Fred Tam, Robert Welsh, Alan Wilman, Yee Hong Yang, Yana Yunusova, Lorne Zinman, for the Canadian ALS Neuroimaging Consortium

## Abstract

**Background:** Amyotrophic lateral sclerosis (ALS) is a disabling and rapidly progressive neurodegenerative disorder. Increasing age is an important risk factor for developing ALS, thus the societal impact of this devastating disease will become more profound as the population ages. A significant hurdle to finding effective treatment has been an inability to accurately quantify cerebral degeneration associated with ALS in humans. Advanced magnetic resonance imaging (MRI) techniques hold promise in providing a set of biomarkers to assist in aiding diagnosis and in efficiently evaluating new drugs to treat ALS.

**Methods:** The Canadian ALS Neuroimaging Consortium (CALSNIC) was founded to develop and evaluate advanced MRI-based biomarkers that delineate biological heterogeneity, track disease progression, and predict survival in a large and heterogeneous sample of ALS patients.

**Findings:** CALSNIC has launched two studies to date (CALSINC-1, CALSNIC-2), acquiring multimodal neuroimaging, neurological, neuropsychological data, and neuropathological data from ALS patients and healthy controls in a prospective and longitudinal fashion from multiple centres in Canada and, more recently, the United States. Clinical and MRI protocols are harmonized across research centres and different MR vendors.

**Interpretation:** CALSNIC provides a multicentre platform for studying ALS biology and developing MRI-based biomarkers.

**Funding:** Canadian Institutes of Health Research, ALS Society of Canada, Brain Canada Foundation, Shelly Mrkonjic Research Fund

## Introduction

Amyotrophic lateral sclerosis (ALS), a type of motor neuron disease (MND), also known as Lou Gehrig’s disease, is the most common motor neuron disorder in adulthood. It can begin at any age but has a peak incidence in the 6th and 7th decades of life; hence the impact of ALS is expected to rise considerably with the aging of the population. The prevalence of ALS is relatively low, due to its rapidly fatal nature: half of those affected die within 3 years of diagnosis, and 90% die within 5 years.^1^

The hallmark pathology of ALS is degeneration of upper motor neurons (UMNs) originating in the motor cortex and of lower motor neurons (LMNs) in the brainstem and spinal cord. However, degeneration often extends beyond the motor regions, involving the frontal and temporal lobes (frontotemporal lobar degeneration, FTLD). The resulting spectrum of behavioural and cognitive manifestations necessitates that ALS be considered as a clinical and biological spectrum of disease rather a singular disease entity.^2^ Motor features include limb weakness, dysarthria, dysphagia, muscle atrophy, fasciculations, and abnormal muscle tone and reflexes. Respiratory failure is the usual cause of death. Cognitive and behavioural dysfunction affects upwards of 50% of patients, with frank dementia occurring in 15% of individuals.^3^

There has been considerable progress in understanding the underlying genetic and biological mechanisms of ALS, yet a fundamental cause of the disease remains elusive.^4^ Approximately 10% of ALS cases have a familial basis. Treatment is largely supportive, with two approved medications in North America that provide modest disease-modifying benefits (riluzole, edaravone). Research activity is brisk in the evaluation of novel therapies in clinical trials. Challenges that continue to stifle progress include i) the absence of a biomarker to quantify response to the therapeutics, and ii) the biological heterogeneity of ALS.

A sensitive, specific, and objective human biomarker of cerebral degeneration would address these two challenges. Magnetic resonance imaging (MRI) is a promising candidate to provide biomarkers because it is objective, non-invasive, imposes minimal risk to the patient, can be readily repeated, and allows regional brain assessment. Conventional structural MRI may reveal hypointensity in the precentral gyral cortex, hyperintensity of the corticospinal tract (CST), and focal atrophy of the motor cortex or frontotemporal regions. However, these changes are qualitative, and neither sensitive nor specific and therefore, clinical MRI remains a tool to rule out disorders that mimic ALS rather than to confirm diagnosis, quantify disease burden or monitor progression.^5^ To obtain useful MRI biomarkers of ALS, something beyond the normal convention is needed.

A remarkable aspect of MRI is its inherent flexibility that enables biological tissues to be depicted with many diverse methods of signal contrast. A growing body of literature supports the potential of several advanced MR methods as biomarkers of ALS progression, survival, and therapeutic effect.^6^ To date, these studies have had limited impact due to several factors. Single centre studies result in biased cohorts with reduced generalizability of the findings to the broader population. A few encouraging studies have made efforts to incorporate multicentre data, however they have been retrospective and lack harmonized protocols.^7,8^ Furthermore, cohorts of matched healthy controls have not been included consistently in studies, and longitudinal datasets are desperately lacking. Sample sizes have been insufficient to allow for robust correlative analysis with clinical subgroups that may reflect biological heterogeneity.

The potential of MRI was well recognized by a group of international academics with an interest in ALS imaging, who met in 2010 to form the Neuroimaging Society in ALS (NiSALS). The meeting led to the publication of consensus guidelines recommending clinical and MRI data that would be desirable in future studies.^9^ This was followed by a NiSALS opinion paper and both papers emphasized the need for multimodal longitudinal MRI conducted at multiple centres using harmonized neuroimaging protocols.^9,10^

In response to these challenges and needs, the Canadian ALS Neuroimaging Consortium (CALSNIC) was founded as a multicentre research platform to facilitate translational research on a national scale. CALSNIC is composed of a multidisciplinary team of scientists with expertise in clinical neuroscience, imaging science, computing science, neuropathology, and biostatistics with a mandate to provide the infrastructure to develop and validate MRI biomarkers in a standardized fashion. The core tenets are a *prospective, systematic, longitudinal*, and *multicentre* approach with clinical and imaging protocols *harmonized* across all centres.

The consortium currently encompasses two studies with a current combined enrollment of over 200 ALS patients and 150 healthy subjects. Both studies follow a schedule of combined clinical and MRI visits at baseline, 4 months, and 8 months:

1. CALSNIC-1 was the initial foundational study that sought to investigate contemporary and multimodal MRI biomarkers. Demographic, clinical, neuropsychological, and multimodal imaging data was collected from participants, with most of them having at least one follow up visit. The study is now closed to enrollment; 87 patients with ALS and related motor neuron diseases and 65 healthy subjects were enrolled across six sites in Canada from 2014 to 2019.
2. CALSNIC-2 operates with a near-identical paradigm to CALSNIC-1. With an opportunity to review CALSNIC-1 and address feedback from site investigators and research staff, the protocol underwent slight but important revisions. Changes were made to the MRI data protocol to address novel questions related to structural and microstructural cerebral changes. A more comprehensive clinical battery was incorporated, and the cognitive assessment became more focused and streamlined. CALSNIC-2 added additional sites in Canada to increase the scope of the consortium and for sample size considerations and began active recruitment in late 2016. In addition, two US partners joined CALSNIC-2 in 2018 to improve generalizability of the study, establish and foster international collaboration, assist with recruitment efforts, and to seek synergy with multi-centre efforts in the US that had a different focus (e.g. biofluid biomarkers).

In what follows, the designs of CALSNIC-1 and CALSNIC-2 (Figure 1) are summarized in greater detail, and their ramifications discussed.

**Figure 1:**
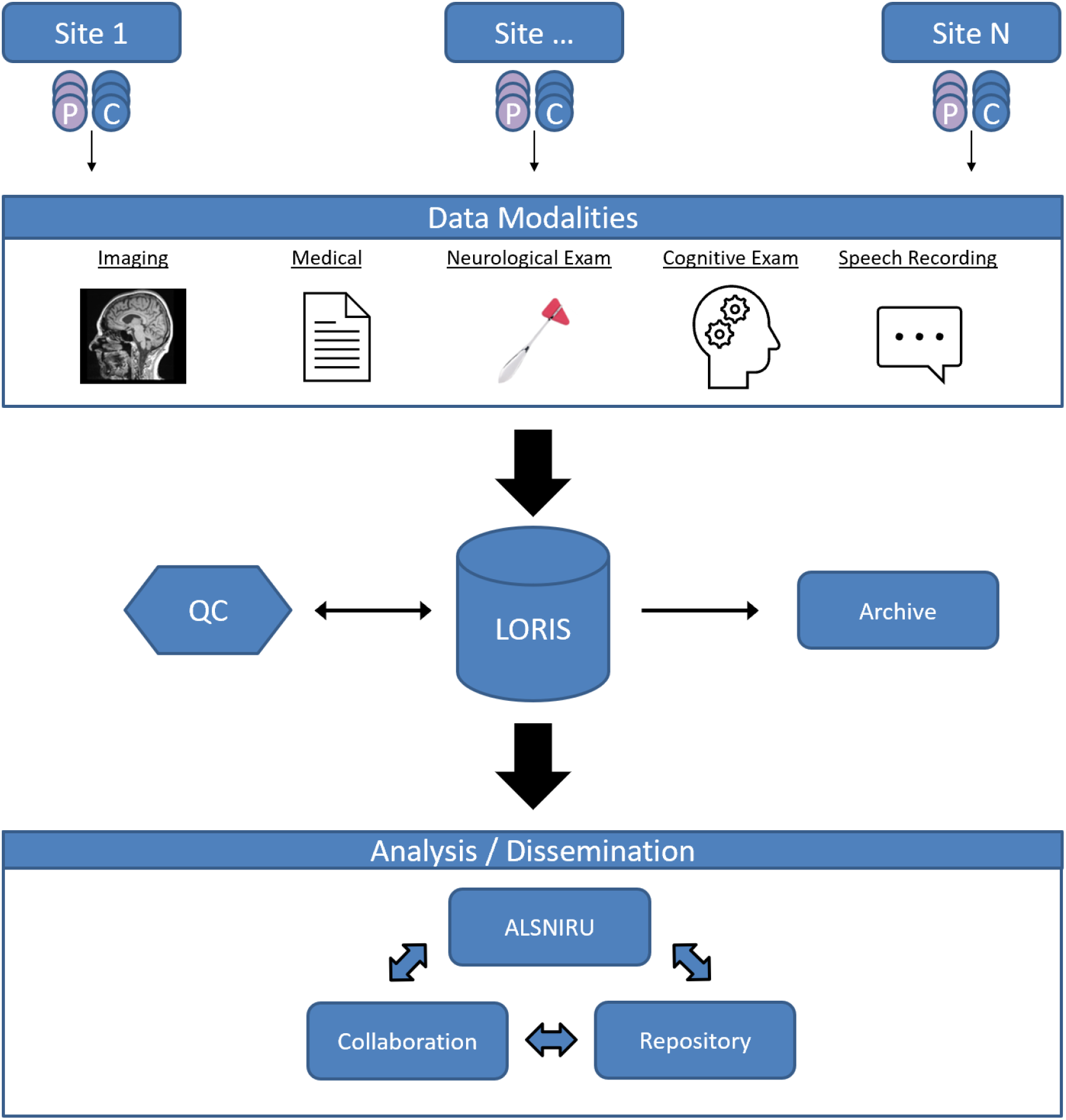
CALSNIC data acquisition and flow. Multimodal data is collected using harmonized protocols across multiple sites. Upload and storage of data is managed using Longitudinal Online Research and Imaging System (LORIS). P=patients with ALS/MND, C=healthy controls, ALSNIRU=ALS Neuroimaging Unit at University of Alberta.

## Methods

### Participants

Patients with ALS and healthy controls were/are recruited from nine North American university centres: University of Alberta in Edmonton, AB (CALSNIC-1, CALSNIC-2); University of British Columbia in Vancouver, BC (CALSNIC-1); University of Calgary in Calgary, AB (CALSNIC-1, CALSNIC-2); Université Laval in Quebec City, QC (CALSNIC-2); University of Miami in Miami, FL (CALSNIC-2); the Montreal Neurological Institute and Hospital in Montreal, QC (CALSNIC-1, CALSNIC-2); Western University in London, ON (CALSNIC-1); University of Toronto in Toronto, ON (CALSNIC-1, CALSNIC-2); and University of Utah in Salt Lake City, UT (CALSNIC-2).

Patients were/are included in CALSNIC-1 and CALSNIC-2 if they are diagnosed with sporadic or familial ALS, and meet the revised El Escorial research criteria^11^ for possible, probable, probable-laboratory supported, or definite ALS. As the projects have been ALS-specific, subjects with a related motor neuron disease (primary lateral sclerosis, progressive muscle atrophy, Kennedy’s disease) or ALS mimics have thus far only been enrolled in smaller numbers. Healthy participants were/are excluded if they have a history of CNS disease *(e.g*., stroke, head injury) or significant psychiatric illness. Results of clinical genetic testing, medical history, family history, and medications are recorded.

Healthy volunteers were recruited as “travelling heads”; each underwent the full MRI protocol at each participating site to acquire data to be used for inter-site and intra-site reliability.

### Clinical Assessments

Assessments are in line with and extend beyond recommendations,^9^ including global measures of disease status: the revised ALS functional rating scale (ALSFRS-R), forced vital capacity (FVC), a neurological exam with a focus on UMN and LMN function, and cognitive and behavioural tests (Table 1). The neurological exam is performed by a neurologist at each visit. The presence of weakness, and UMN and LMN signs were recorded in a binary fashion (presence or absence of weakness, hyperreflexia, spasticity, etc.) in CALSNIC-1. For CALSNIC-2, the examination was expanded for detailed scoring.

**Table 1:**
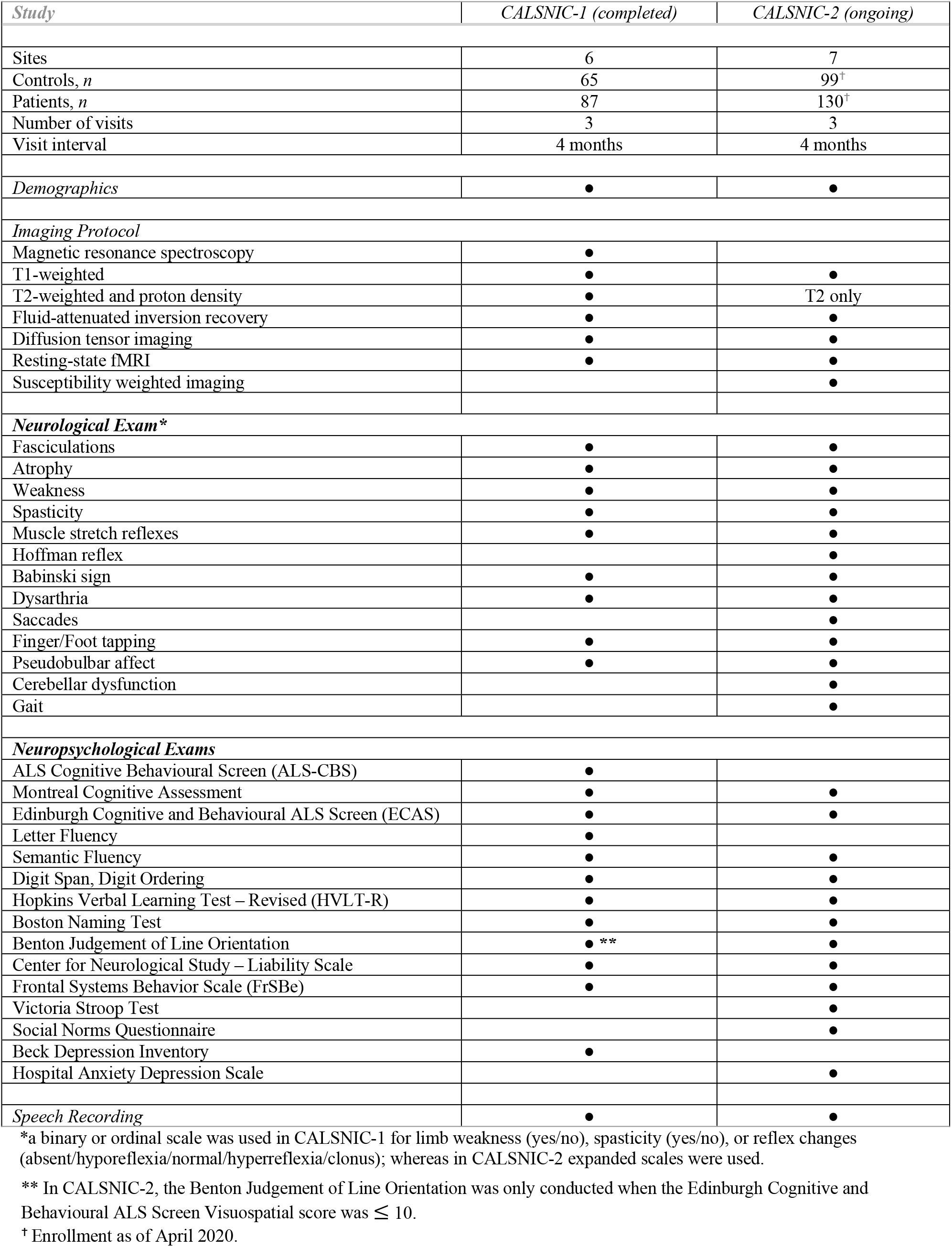
Overview of CALSNIC studies.

The protocol for cognitive and behavioural testing follows published guidelines for ALS.^2^ The Edinburgh Cognitive and Behavioral ALS Screen (ECAS)^12^, a brief validated screening assessment designed specifically for this patient population, was implemented in both studies and at each visit in CALSNIC-2 upon the release of its alternate version. To increase sensitivity and assess for co-morbid pathologies, the ECAS is supplemented with additional tests of executive functioning, letter and semantic fluency, attention, memory, language, and visuospatial function. A French-Canadian version of the ECAS did not exist at the start of the project. Through the existing partnership in CALSNIC with researchers at the Montreal and Quebec City sites, the instrument was formally translated, and normative healthy control data were collected. Existing French versions of other cognitive tests are used where available. Furthermore, there are efforts underway to develop North American norms for the English ECAS.

Participants further undergo speech evaluation to study presence and severity of bulbar motor deficits. This involves recording of speech with three tasks: Speech Intelligibility Test (SIT)^13^ during which speech intelligibility and speaking rate are assessed, Maximum Phonation Task, during which voice acoustics and quality are evaluated, and Passage Reading (Bamboo passage),^14^ to obtain speech and pause duration statistics.

A standardized post-mortem examination is available to patients in CALSNIC-1. The brains and spinal cords of ALS patients who have consented to autopsy and have subsequently passed away were processed by the Neuropathology team in the patient’s home institution following a standardized blocking and staining protocol, and formalin fixed paraffin embedded blocks from 9 standardized regions of interest were sent for central Neuropathology review. Quantitative and semi-quantitative histologic analysis of this central pathology review is currently underway. Neuropathologic findings will be compared to MRI data.

### Imaging Protocols

CALSNIC-1 and CALSNIC-2 participants underwent/undergo a standardized one-hour MRI protocol (Figure 2) at the 3 time-points (baseline, 4 months, and 8 months). For a detailed outline of the protocol and sequence parameters for each project, refer to Table 2.

**Figure 2:**
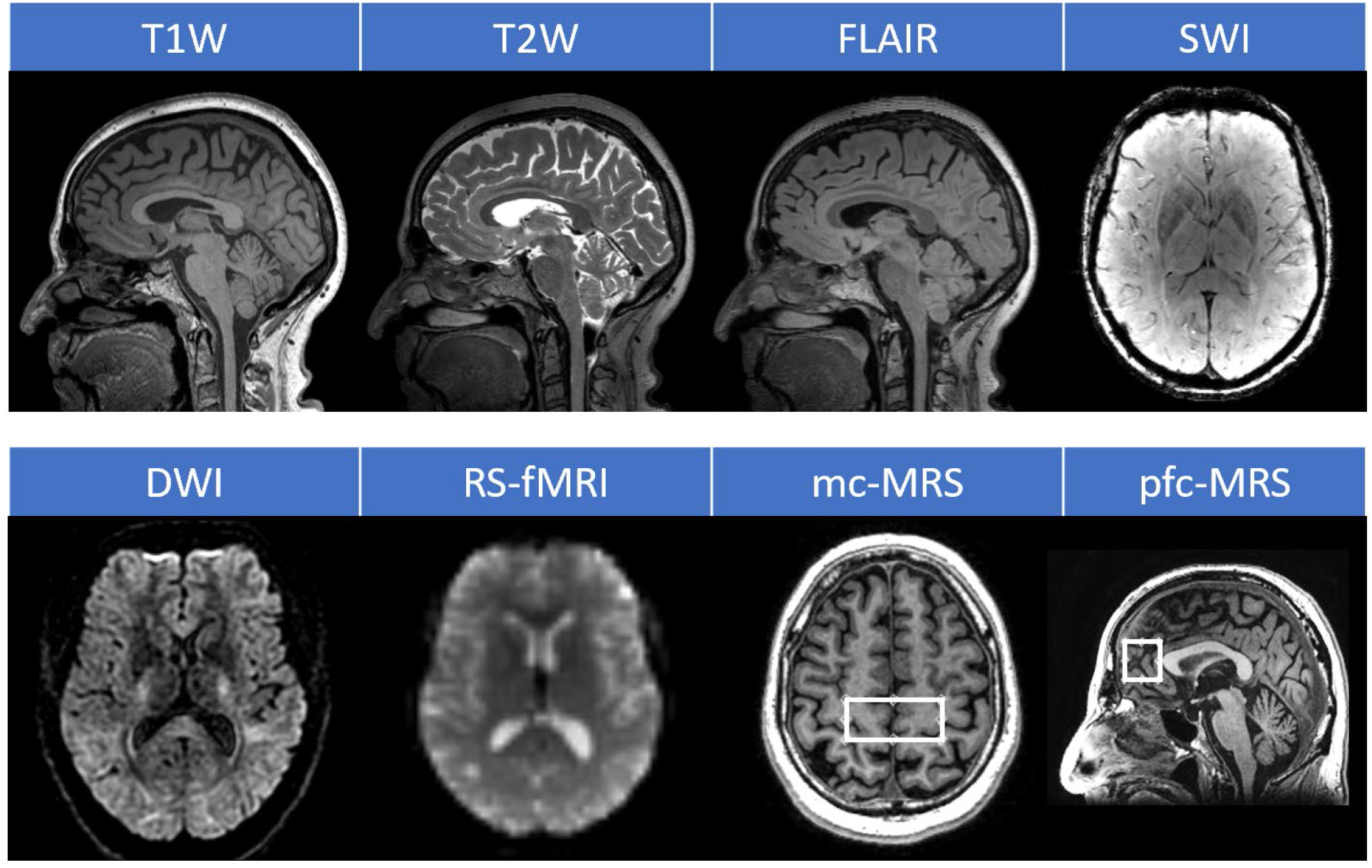
Representative images from the CALSNIC MRI imaging protocol. T1W=T1-weighted, T2W=T2-weighted, FLAIR=Fluid Attenuated Inversion Recovery, SWI=Susceptibility Weighted Imaging (CALSNIC-2), DWI=Diffusion Weighted Imaging, RS-fMRI=Resting State Functional MRI, MRS=Magnetic Resonance Spectroscopy (CALSNIC-1) showing voxel placement in the motor cortex (mc) and prefrontal cortex (pfc) from where data is acquired.

**Table 2:**
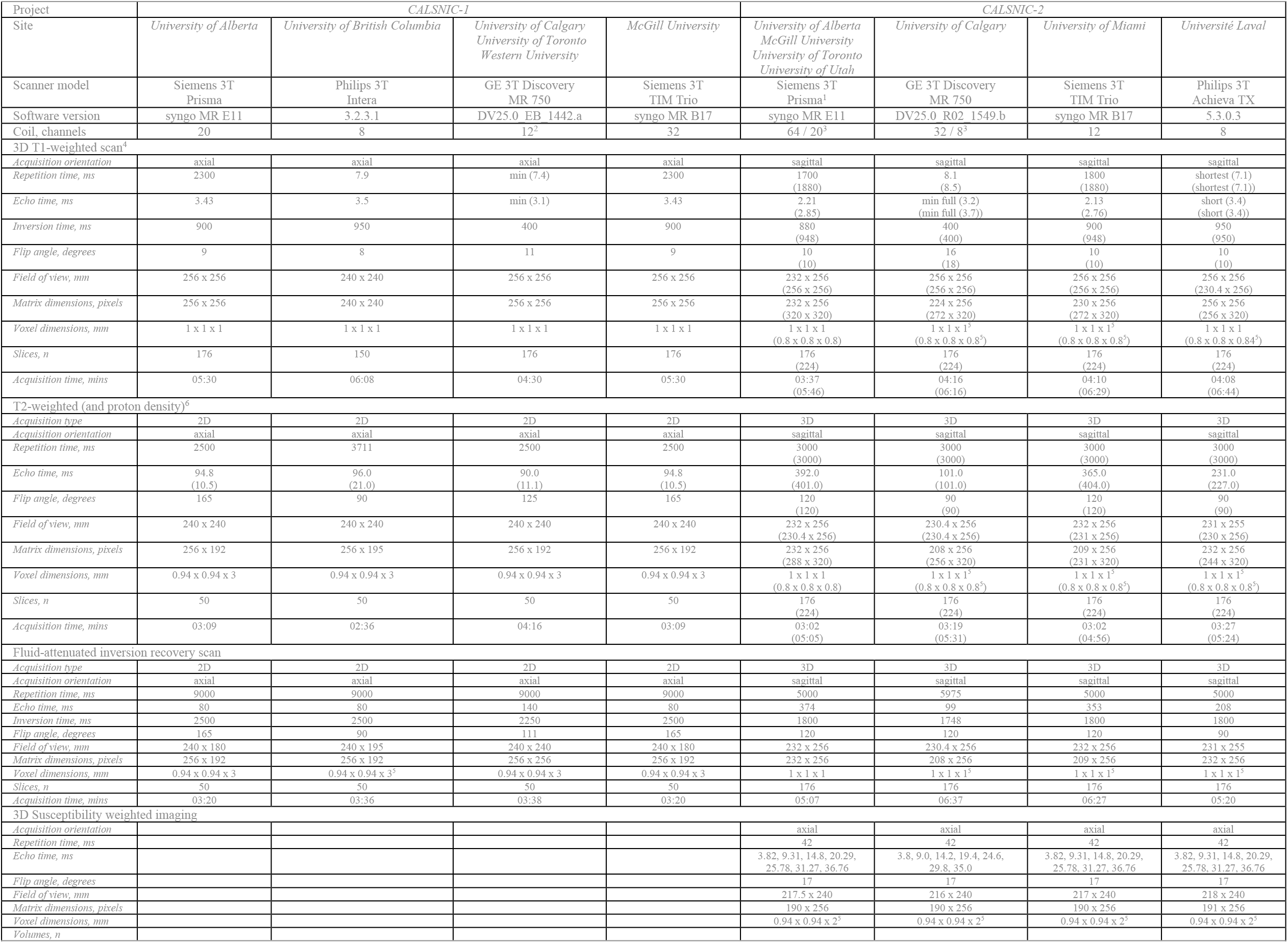
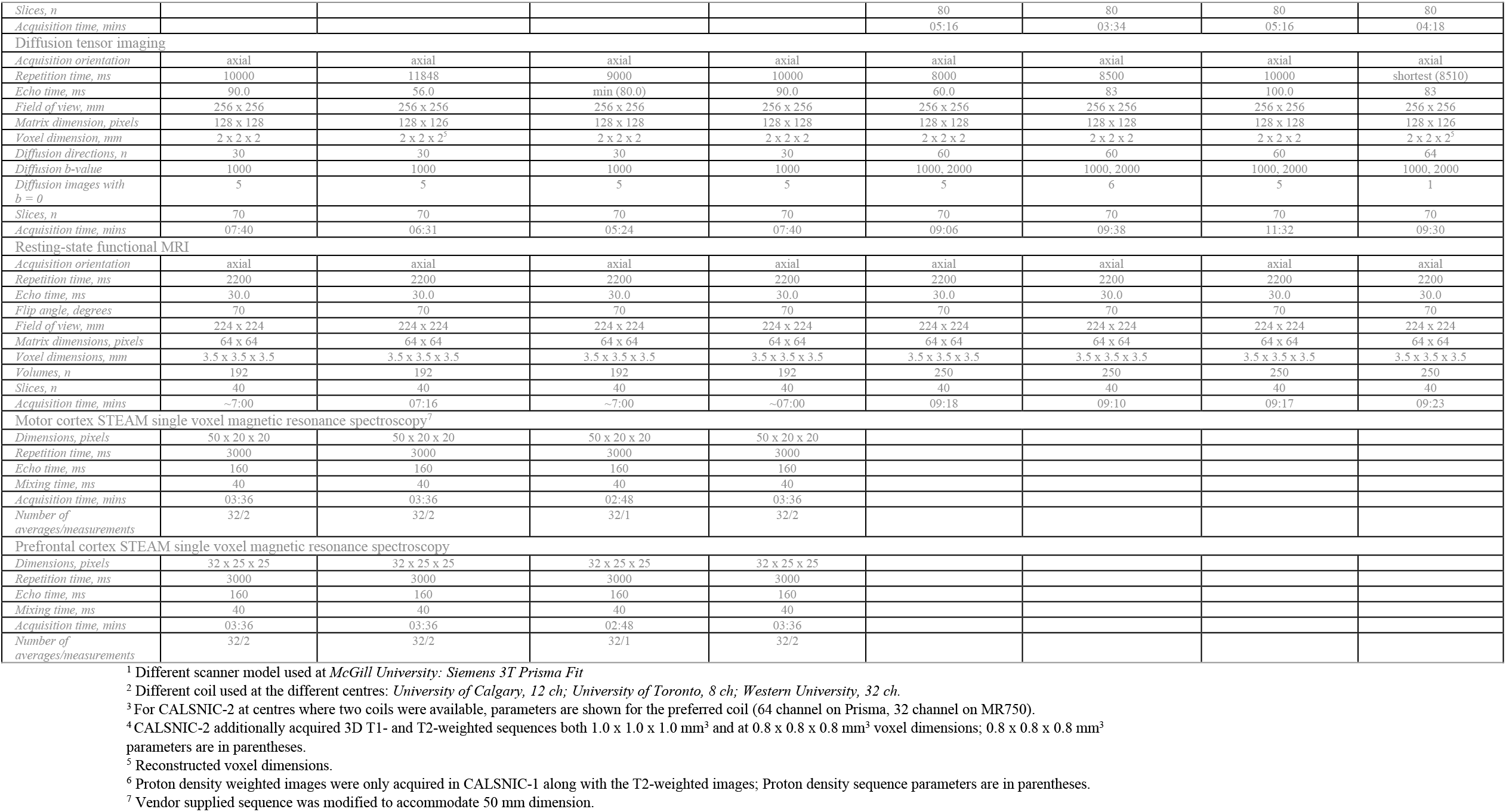
CALSNIC MR Protocol.

#### Overview of CALSNIC imaging protocols

The two protocols have some common elements. High resolution anatomical sequences, such as three-dimensional (3D) T1-weighted (T1W) and T2-weighted (T2W), for assessment of grey matter (GM) density, cortical thickness, image texture, and coincidental pathologies such as stroke, tumor, or developmental anomalies. Diffusion tensor imaging (DTI) assesses fractional anisotropy (FA), mean diffusivity (MD), structural connectivity, and other indices of white matter (WM) integrity. Resting-state functional MRI (rs-fMRI) assesses functional connectivity. CALSNIC-1 includes single voxel magnetic resonance spectroscopy (MRS) for assessment of neuronal integrity and gliosis in the motor and prefrontal cortices;^15^ this was not included in CALSNIC-2 due to time limitations. Instead, CALSNIC-2 added susceptibility weighted imaging (SWI) to quantify cerebral iron deposition, a second *b*-value of diffusion weighting to allow for diffusion kurtosis imaging, and multi-resolution 3D anatomical imaging.

#### General approach to standardization of MR Imaging

A multi-faceted approach was taken to optimize inter-site data comparability: 1) use of only 3 T MRI systems, 2) use of existing, vendor-supplied, sequences on MRI systems (rather than attempting to develop and implement new sequences), 3) iterative adjustment of sequence parameters for each vendor (GE Healthcare, Siemens Healthineers, Philips Healthcare) to provide comparable data with qualitative and/or quantitative assessments of blur, distortion, signal-to-noise ratio, homogeneity, and artifacts, and 4) the use of fixed spatial parameters (image resolution, slice thickness, number of slices, field of view) across vendors whenever possible. Inter-vendor differences existed for head receiver coil arrays; a 64-channel array was preferred where available due to its preferable signal-to-noise ratio (SNR) characteristics. As some subjects would not fit into the 64-channel coil, the MR protocol includes sequences for the sites’ alternate coil array *(e.g*., 20-channel head coil).

#### T1W

Beyond visual inspection, 3D T1W MRI data can be further post-processed to quantify disease burden objectively by various methods, yielding several candidate imaging-based biomarkers. For example, previous voxel-based morphometry (VBM) studies in ALS have revealed focal atrophy of the GM in the motor and frontotemporal areas,^16^ and subcortical GM including the basal ganglia and thalamus.^17^ Longitudinal decline of GM density in the motor cortex has been detected over a period of six months,^18^ with additional involvement of the GM in the premotor, basal ganglia and frontotemporal regions after nine months.^19^ Surface-based morphometry (SBM) has revealed cortical thinning localized to the motor cortex.^20^ More recently, 3D texture analysis in ALS has revealed global alterations of grey and white matter,^21^ including the CST,^22^ and motor cortex and subcortical structures.^23^

The CALSNIC-1 dataset was subjected to deformation-based morphometry (DBM) analysis to study longitudinal changes in GM and WM, revealing progressing atrophy in the precentral gyrus and frontal and parietal WM.^24^

The CALSNIC-1 and CALSNIC-2 T1W protocol is acquired at 1 mm isotropic spatial resolution to provide seamless harmonization and integration across the projects. CALSNIC-2 additionally acquires 3D T1W MRI data at 0.8 mm isotropic resolution to study the impact of multi-resolution images on post-processing analytics. For specific details on the 3D T1W sequence parameters, refer to Table 2.

#### T2W/PD

Signal intensity changes seen on T2W, fluid-attenuated inversion recovery (FLAIR) and proton density (PD)-weighted sequences are not sensitive or specific in ALS,^5^ although their presence may be reflective of different pathologies and may assist in differential diagnosis. These sequences were included to assess for co-existing disease and to be available for image processing pipelines (e.g., skull stripping) that some investigators may employ.

#### DTI

DTI has been the gold standard for detecting microstructural WM alterations and has consistently revealed abnormal axonal integrity by an increased MD, a measure of bulk diffusion, and decreased FA, an indirect measure of structural integrity, in patients affected by ALS. Studies have demonstrated changes in the CST and voxel-wise whole brain analyses have further revealed changes beyond the motor system, including in frontotemporal areas,^25^ and in the corpus callosum (CC).^26^

The CALSNIC-1 and CALSNIC-2 DTI protocol is acquired at 2 mm isotropic resolution with 30 non-collinear directions of diffusion weighting *(b* = 1000 s/mm^2^) and 5 volumes of *b* = 0 s/mm^2^. CALSNIC-2 additionally collects 30 volumes at *b* = 2000 s/mm^2^. Refer to Table 2 for the whole brain DTI sequence parameters.

A longitudinal analysis of the CALSNIC-1 DTI dataset incorporating corrective approaches to site variance revealed progressive decline in WM integrity in the CST and frontal lobes.^27^

#### SWI

The SWI sequence exploits tissue magnetic susceptibility differences to generate unique and enhanced contrast images such as R2* and quantitative susceptibility maps (QSM), providing an objective measure of cerebral iron accumulation. Past SWI studies in ALS have revealed signal abnormalities suggesting abnormal iron metabolism or deposition diffusely in WM^28^ and the motor cortex.^29^ Texture analysis applied to SWI images has demonstrated alterations within the precentral gyrus and basal ganglia.^30^

3D SWI sequences were only acquired as part of the CALSNIC-2 imaging protocol. SWI parameters are outlined in Table 2.

#### MRS

MRS permits objective and non-invasive measurements of cerebral chemicals, including markers of neuronal integrity (N-acetylaspartate, NAA), astrogliosis (myo-inositol, mI), and neurotransmitter metabolism such as glutamate and GABA. Aberrations in these and other metabolites have been quantified in ALS in motor and extra-motor cerebral regions, brainstem and spinal cord.^31^

The MRS protocol in CALSNIC-1 aimed to assess NAA and mI in the primary motor cortex and mesial prefrontal cortex. A single voxel stimulated echo acquisition mode (STEAM) sequence was used that was optimized for detection of these two metabolites.^32,33^ The mesial prefrontal cortex was chosen as it is pathologically involved in ALS.^34^ Prior literature has also demonstrated superior discriminatory power of NAA/mI ratio both in the motor^32^ and prefrontal^33^ cortices. The positioning of each voxel of interest (VOI) was carefully chosen to ensure consistent landmarking of the VOI across subjects and centres.

The baseline multicentre MRS from CALSNIC-1 has revealed reduced metabolite concentration ratios in the motor cortex of ALS patients.^15^

#### rs-fMRI

Resting-state fMRI measures fluctuations in the blood oxygen level-dependent (BOLD) signal in the brain when the participant is not occupied with any task-based stimuli; thus identifying potential abnormal functioning of resting-state networks (RSNs). In ALS, the sensorimotor network (SMN) and default mode network (DMN) have altered functional connectivity in both motor and non-motor regions.^35–37^

Resting-state fMRI was/is acquired in CALSNIC-1 and CALSNIC-2 at a 3.5 mm isotropic resolution. CALSNIC-1 collected 192 volumes in 7 minutes, and the acquisition time is increased in CALSNIC-2 to 10 minutes to acquire 250 volumes. Refer to Table 2 for further details on the rs-fMRI parameters.

### Data storage

CALSNIC-1 acquired and stored clinical data through the Research Electronic Data Capture^38^ (REDCap; https://www.project-redcap.org/) platform. REDCap is a secure web application that provides researchers the functionality to collect and manage various types of data across multiple centres. Neuroimaging data for CALSNIC-1 was uploaded by each site through a file transfer protocol (FTP) server as compressed Digital Imaging and Communications in Medicine (DICOM) images. The REDCAP application and FTP server were deployed at the University of Calgary under the management of the Canadian Neuromuscular Disease Registry.^39^

For CALSNIC-2, the consortium transitioned to the Longitudinal Online Research and Imaging System^40^ (LORIS; http://loris.ca/) to store both clinical and imaging data on a unified platform. This online neuroimaging software is open-source, and provides a secure framework to store multimodal data, including clinical and neuroimaging data, for longitudinal multicentred projects. The LORIS instance for CALSNIC-2 is hosted at the University of Alberta.

Access to clinical and imaging data is granted through secure and restricted online portals to research personnel. Each member of the project requires a username and password and has a unique set of permissions assigned to access, upload, and audit data.

### Site training

Research personnel are trained online and in-person on study procedures including clinical assessments, neuro-psychometric testing, imaging acquisition, and electronic data capture through the database web interface to ensure standardized data collection and storage across all sites. In particular, the placement of the VOI in MRS acquisitions (CALSNIC-1 only) was specifically reviewed with MR technologists to ensure consistency and validity of the data. Monthly teleconference calls are held with research assistants from the participating centres to address site-specific issues and to provide any study related updates. Training sessions are held if staffing resources change, or if quality control issues are identified.

### Quality control and reliability

#### Clinical data

Clinical data validation and quality checking is multilayered with the initial check enforced at the time of data entry. This is accomplished by restricting database fields to be associated with specific data types (numerical, date, string, etc.), to have defined numerical ranges, and to be automatically calculated, if applicable. The data entered must adhere to such specifications and any invalid input must be resolved prior to saving the clinical data in the database to ensure a syntactical level of data validation. Furthermore, data auditing is conducted every three months across the entire clinical dataset to provide a semantic level of data validation.

#### Data file naming conventions

Behavioural and imaging files adhered to naming specifications. Participants are assigned a de-identifying ID prior to acquiring and uploading data. CALSNIC-1 naming convention consisted of a concatenation of the site name *(e.g*., Edmonton), control or patient number (e.g., C001 or P001), and the visit label (e.g., month0, month4, month8) (e.g., Edmonton_C001_month0). To comply with the LORIS system specifications, the CALSNIC-2 naming convention was modified. It consisted of a concatenation of a three-letter site abbreviation and a three-digit participant number (e.g. EDM001), a six-digit randomly generated unique identification number *(e.g*., 123456), and a visit label *(e.g*., V1, V2, V3) resulting in the label EDM001_123456_V1.

#### Review for image quality, artifacts and medical co-morbidities

An automated quality control pipeline was designed and set-up to validate the incoming MRI datasets. This involves: 1) verification of file name; 2) ensuring no patient identifying information is present within the file;3) verification of acquisition parameters against a reference protocol for each MRI system and imaging sequence. If any of these conditions are violated or if there is a divergence in the acquisition parameters of the uploaded sequences against the reference protocol, then the user is notified to investigate the issue, and if possible, to clean and re-upload the dataset. The MRI data are then screened for medical co-morbidities at the first time point of data collection (0 months) and escalated as necessary to review by a neuroradiologist. Structural abnormalities and artifacts are noted, and the imaging session is rescheduled if necessary.

#### Reliability: Assessment of site difference

The Alzheimer’s Disease Neuroimaging Initiative (ADNI) phantom (Magphan EMR051, The Phantom Laboratory, Salem, NY) was initially used at all sites prior to subject enrollment to establish baseline MRI system characteristics and to calibrate them for geometric uniformity, SNR and CNR. To determine intra- and inter-site variability in the MRI measures obtained, six healthy human phantoms (“travelling heads”) for CALSNIC-1 and ten for CALSNIC-2 underwent the complete imaging protocol twice at each participating centre. The time interval chosen between the two scans was a few hours to consider the effect of day-to-day physiological variations, assuming that longer term variations would be addressed by a combination of local quality control procedures and preventative maintenance procedures established at each site. The travelling head images are available to derive scaling and corrective factors to transform data from sites into a common data space prior to analysis, and to assess inter-site and intra-site variance. The travelling head data was used for reliability assessments for MRS,^15^ DTI,^27^ and texture^41^ modalities.

## Discussion

CALSNIC is a clinical investigative platform for neuroimaging-focused translational research. It has the capacity to collect data in a prospective and longitudinal manner using harmonized protocols across sites. Two core studies have operated within the consortium; enrollment is complete for CALSNIC-1 and is ongoing for CALSNIC-2. An important rationale for the multicentre design is that biomarkers must eventually be able to perform in the multicentre drug trial arena as outcome measures. At its inception, sites were chosen based on the requirement of a multidisciplinary ALS clinic with research experience and the presence of an active MRI research centre with appropriate technical expertise.

The Canadian ALS clinical and research community is relatively small with investigators well known to each other, making the operation of multicentre studies easier in many respects. As evidence of this, the community has a strong national clinical trials research network (Canadian ALS Research Network) and a national registry (Canadian Neuromuscular Disease Registry),^39^ with which CALSNIC interfaces. CALSNIC has increased research capacity and throughput at these sites, including the introduction of ALS imaging research at many.

CALSNIC sites are in both predominantly English or French speaking communities, with clinical research protocols provided in both languages. There is geographic representation, from Vancouver in the west to as far east as Quebec City. Recent expansion to include two US sites and implementation of Spanish instruments where available have further enriched the study. Other international endeavours include academic collaborations,^27^ and the deposition of data into the Jena MRI repository^42^ of NiSALS.

CALSNIC publications are forthcoming that report results from multicentre MRS,^15^ DTI,^27^ structural,^24^ texture,^41,43^, and neuropathology data. The richness and extensiveness of the data provides a unique opportunity for advanced analytical methods (multivariate classifiers, AI) to help discriminate biological variability. The initiative will undoubtedly evolve over time and has the capacity to meet individual and collective research objectives. This could include the incorporation of other measures of cerebral degeneration such as PET imaging, transcranial magnetic stimulation, and advanced measures of LMN dysfunction (e.g. motor unit number estimation). A necessary evolution of CALSNIC will be to address the limitation of the lack of biofluid sampling (serum, cerebrospinal fluid); this is indeed under active development by lead investigators.

## Conclusion

CALSNIC is an interdisciplinary venture aimed at validating promising MRI biomarkers and to investigate biological heterogeneity in ALS patients. A better understanding of the structural, chemical, and functional alterations associated with different phenotypes of ALS (e.g., disease progression rate, survival, cognitive impairment) may translate back to the bench with newly realized questions for researchers. A validated and objective imaging biomarker would facilitate the evaluation of novel therapeutics in phase II trials with subject selection and outcome measures that have better sensitivity to disease progression than existing clinical outcome measures. This would translate to increased trial efficiency with reduced sample sizes, trial duration, cost and ultimately the timelier discovery of effective therapy. CALSNIC has created opportunities for addressing research questions at an international level.

## Data Availability

Details of the protocols in this manuscript will be made available at http://calsnic.org/.

## Notes

### Competing Interest Statement

The authors have declared no competing interest.

### Clinical Trial

NCT02405182 and NCT03362658

### Clinical Protocols

http://calsnic.org/

### Funding Statement

CALSNIC is funded by the Canadian Institutes of Health Research, the ALS Society of Canada, the Brain Canada Foundation, and the Shelly Mrkjonjic Research Fund.

### Author Declarations

The study was approved by the health research ethics boards of each participating site, and all participants gave written informed consent.

### Summary of Updates

Author list updated. Minor clarifications to Methodology section. Tables 1 and 2 updated.

## References

1 del Aguila MA, Longstreth WT, McGuire V, Koepsell TD, van Belle G. Prognosis in amyotrophic lateral sclerosis: A population-based study. Neurology 2003; 60: 813–9.

2 Strong MJ, Grace GM, Freedman M, et al. Consensus criteria for the diagnosis of frontotemporal cognitive and behavioural syndromes in amyotrophic lateral sclerosis. Amyotroph Lateral Scler 2009;10:131–46.

3 Lomen-Hoerth C, Murphy J, Langmore S, Kramer JH, Olney RK, Miller B. Are amyotrophic lateral sclerosis patients cognitively normal? Neurology 2003; 60: 1094–7.

4 Taylor JP, Brown RH, Cleveland DW. Decoding ALS: From genes to mechanism. Nature. 2016; 539:197–206.

5 Filippi M, Agosta F, Abrahams S, et al. EFNS guidelines on the use of neuroimaging in the management of motor neuron diseases. Eur J Neurol 2010; 17: 526–e20.

6 Menke RAL, Agosta F, Grosskreutz J, Filippi M, Turner MR. Neuroimaging Endpoints in Amyotrophic Lateral Sclerosis. Neurotherapeutics 2017; 14: 11–23.

7 Müller H-P, Turner MR, Grosskreutz J, et al. A large-scale multicentre cerebral diffusion tensor imaging study in amyotrophic lateral sclerosis. J Neurol Neurosurg Psychiatry 2016; 87: 570–9.

8 Müller HP, Agosta F, Riva N, et al. Fast progressive lower motor neuron disease is an ALS variant: A two-centre tract of interest-based MRI data analysis. NeuroImage Clin 2018; 17: 145–52.

9 Turner MR, Grosskreutz J, Kassubek J, et al. Towards a neuroimaging biomarker for amyotrophic lateral sclerosis. Lancet Neurol 2011; 10: 400–3.

10 Filippi M, Agosta F, Grosskreutz J, et al. Progress towards a neuroimaging biomarker for amyotrophic lateral sclerosis. Lancet Neurol. 2015; 14: 786–8.

11 Brooks BR, Miller RG, Swash M, Munsat TL. El Escorial revisited: Revised criteria for the diagnosis of amyotrophic lateral sclerosis. Amyotroph Lateral Scler Other Mot Neuron Disord 2000; 1: 293–9.

12 Abrahams S, Newton J, Niven E,… JF-… lateral sclerosis and, 2014 undefined. Screening for cognition and behaviour changes in ALS. Taylor Fr https://www.tandfonline.com/doi/abs/10.3109/21678421.2013.805784 (accessed Oct 23, 2019).

13 Beukelman D. Speech Intelligibility Test for Windows. 1996 https://www.researchgate.net/publication/260387243 (accessed June 19, 2020).

14 Barnett C, Green JR, Marzouqah R, et al. Reliability and validity of speech & pause measures during passage reading in ALS. Amyotroph Lateral Scler Front Degener 2020; 21: 42–50.

15 Srivastava O, Hanstock C, Chenji S, et al. Cerebral degeneration in amyotrophic lateral sclerosis. Neurol Clin Pract 2019; 9: 400–7.

16 Senda J, Kato S, Kaga T, et al. Progressive and widespread brain damage in ALS: MRI voxel-based morphometry and diffusion tensor imaging study. Amyotroph Lateral Scler 2011; 12: 59–69.

17 Bede P, Elamin M, Byrne S, et al. Basal ganglia involvement in amyotrophic lateral sclerosis. Neurology 2013; 81: 2107–15.

18 Unrath A, Ludolph AC, Kassubek J. Brain metabolites in definite amyotrophic lateral sclerosis. J Neurol 2007; 254: 1099–106.

19 Agosta F, Gorno-Tempini ML, Pagani E, et al. Longitudinal assessment of grey matter contraction in amyotrophic lateral sclerosis: A tensor based morphometry study. Amyotroph Lateral Scler 2009;10:168–74.

20 Verstraete E, Veldink JH, Hendrikse J, Schelhaas HJ, van den Heuvel MP, van den Berg LH. Structural MRI reveals cortical thinning in amyotrophic lateral sclerosis. J Neurol Neurosurg Psychiatry 2012; 83: 383–8.

21 Maani R, Yang YH, Emery D, Kalra S. Cerebral degeneration in amyotrophic lateral sclerosis revealed by 3-dimensional texture analysis. Front Neurosci 2016; 10. DOI: 10.3389/fnins.2016.00120

22 Ishaque A, Mah D, Seres P, et al. Corticospinal tract degeneration in ALS unmasked in T1-weighted images using texture analysis. Hum Brain Mapp 2019; 40: 1174–83.

23 Ishaque A, Mah D, Seres P, et al. Evaluating the cerebral correlates of survival in amyotrophic lateral sclerosis. Ann Clin Transl Neurol 2018; 5: 1350–61.

24 Dadar M, Manera AL, Zinman L, et al. Cerebral atrophy in amyotrophic lateral sclerosis parallels the pathological distribution of TDP43. Brain Commun DOI:10.1093/BRAINCOMMS/FCAA061.

25 Agosta F, Pagani E, Petrolini M, et al. Assessment of White Matter Tract Damage in Patients with Amyotrophic Lateral Sclerosis: A Diffusion Tensor MR Imaging Tractography Study: Fig 1. Am J Neuroradiol 2010; 31: 1457–61.

26 Filippini N, Douaud G, Mackay CE, Knight S, Talbot K, Turner MR. Corpus callosum involvement is a consistent feature of amyotrophic lateral sclerosis. Neurology 2010; 75: 1645 LP – 1652.

27 Kalra S, Müller H-P, Ishaque A, et al. A prospective harmonized multicentre DTI study of cerebral white matter degeneration in ALS. Neurology 2020.

28 Prell T, Hartung V, Tietz F, et al. Susceptibility-Weighted Imaging Provides Insight into White Matter Damage in Amyotrophic Lateral Sclerosis. PLoS One 2015; 10: e0131114.

29 Yu J, Qi F, Wang N, et al. Increased iron level in motor cortex of amyotrophic lateral sclerosis patients: An in vivo MR study. Amyotroph Lateral Scler Front Degener 2014; 15: 357–61.

30 Johns SLM, Ishaque A, Khan M, Yang Y-H, Wilman AH, Kalra S. Quantifying changes on susceptibility weighted images in amyotrophic lateral sclerosis using MRI texture analysis. Amyotroph Lateral Scler Front Degener 2019; 20: 396–403.

31 Kalra S. Magnetic Resonance Spectroscopy in ALS. Front Neurol 2019; 10: 482.

32 Kalra S, Hanstock CC, Martin WRW, Allen PS, Johnston WS. Detection of Cerebral Degeneration in Amyotrophic Lateral Sclerosis Using High-Field Magnetic Resonance Spectroscopy. Arch Neurol 2006; 63: 1144.

33 Usman U, Choi C, Camicioli R, et al. Mesial Prefrontal Cortex Degeneration in Amyotrophic Lateral Sclerosis: A High-Field Proton MR Spectroscopy Study. Am J Neuroradiol 2011; 32: 1677 LP – 1680.

34 Brettschneider J, Del Tredici K, Toledo JB, et al. Stages of pTDP-43 pathology in amyotrophic lateral sclerosis. Ann Neurol 2013; 74: 20–38.

35 Welsh RC, Jelsone-Swain LM, Foerster BR. The utility of independent component analysis and machine learning in the identification of the amyotrophic lateral sclerosis diseased brain. Front Hum Neurosci 2013; 7. DOI:10.3389/fnhum.2013.00251.

36 Chenji S, Jha S, Lee D, et al. Investigating Default Mode and Sensorimotor Network Connectivity in Amyotrophic Lateral Sclerosis. PLoS One 2016; 11: e0157443.

37 Schulthess I, Gorges M, Müller H-P, et al. Functional connectivity changes resemble patterns of pTDP-43 pathology in amyotrophic lateral sclerosis. Sci Rep 2016; 6: 38391.

38 Harris PA, Taylor R, Thielke R, Payne J, Gonzalez N, Conde JG. Research electronic data capture (REDCap)—A metadata-driven methodology and workflow process for providing translational research informatics support. J Biomed Inform 2009; 42: 377–81.

39 Korngut L, Genge A, Johnston M, et al. Establishing a Canadian registry of patients with amyotrophic lateral sclerosis. Can J Neurol Sci 2013; 40: 29–35.

40 Das S, Zijdenbos AP, Harlap J, Vins D, Evans AC. LORIS: a web-based data management system for multi-center studies. Front Neuroinform 2012; 5. DOI:10.3389/fninf.2011.00037.

41 Ta D, Khan M, Ishaque A, et al. Reliability of 3D texture analysis: A multicenter MRI study of the brain. J Magn Reson Imaging 2019;: jmri.26904.

42 Steinbach R, Gaur N, Stubendorff B, Witte OW, Grosskreutz J. Developing a Neuroimaging Biomarker for Amyotrophic Lateral Sclerosis: Multi-Center Data Sharing and the Road to a “Global Cohort”. Front Neurol 2018; 9: 1055.

43 E Elahi GMM, Kalra S, Zinman L, Genge A, Korngut L, Yang YH. Texture classification of MR images of the brain in ALS using M-CoHOG: A multi-center study. Comput Med Imaging Graph 2020; 79:101659.

